# The influence of sidedness in unilateral cleft lip and palate on mid facial growth at 5 years of age

**DOI:** 10.1101/2023.12.05.23299328

**Authors:** Matthew Fell, David Chong, Paras Parmar, Ting-Li Su, Lars Enocson, Bruce Richard

## Abstract

**Objective:** To determine whether facial growth at five years is different for children with a right versus left sided cleft lip and palate.

**Design:** Retrospective cohort study

**Setting:** Nine UK cleft centres

**Patients:** Patients born between 2000-2014 with a complete unilateral cleft lip and palate (UCLP)

**Main outcomes measure:** 5-Year-Old’s Index scores

**Results:** 378 children were included. 122 (32%) had a right sided UCLP and 256 (68%) had a left sided UCLP. 5-Year-Old’s index scores ranged from 1 (good) to 5 (poor). There was a higher proportion of patients getting good scores (1 and 2) in left UCLP (43%) compared to right UCLP (37%) but there was weak evidence for a difference (Adjusted summary odds ratio 1.27, 95% CI 0.87 to 1.87; P=0.22).

**Conclusions:** Whilst maxillary growth may be different for left versus right sided UCLP, definitive analysis requires older growth indices and arch forms.

## Background

There is directional asymmetry in the sidedness of unilateral cleft lip and palate (UCLP) with left sided UCLP twice as prevalent as right sided UCLP.^1^ Anatomical differences have been noted in the soft tissues of the face when orofacial cleft cohorts are stratified by sidedness; right sided unilateral cleft lip with or without palate (UCL/P) has been associated with a greater degree of hypoplasia of the lateral lip element^2^, more pronounced facial disfigurement^3,4^ and facial asymmetry^5^ compared to left sided UCL/P. As these differences have been observed in the soft tissues of the face between left and right sided clefts, it is plausible to consider that there could be differences in the underlying bony structures of the facial skeleton.

Maxillary growth is an important outcome in patients born with UCLP due to the risk of maxillary hypoplasia and subsequent retrusion, which results in malocclusion and malalignment of the jaws, lips and tongue.^6,7^ Most maxillary growth occurs in the first eight years of life from ossification centres located in the transverse palatal suture, giving rise to anterior-posterior growth and the mid-palatal suture, giving rise to transverse growth.^8,9^ Many studies have focused on the impact of extrinsic factors on midfacial growth and it is widely accepted that surgical intervention on the palate can harm maxillary growth plates.^9–13^ Furthermore, reports from rare cohorts of patients with unrepaired orofacial clefts have demonstrated intrinsic growth deficiency compared to controls.^10,14^ Less is known about the impact of intrinsic factors for maxillary growth such as cleft phenotype and sex.

The aim of this study is to investigate the relationship between the sidedness of UCLP and facial growth at 5 years of age.

## Methods

### Registration

This study was nested within a larger ongoing investigation of midfacial growth in children born with complete UCLP (IRAS: 225282, Research Registry UIN: 4068). The purpose of the larger study is to investigate intrinsic factors leading to poor mid facial growth via the analysis of pre-surgical 3D scans of the maxillary arch.

### Participants and exposure

Children were included if they were born in the United Kingdom (UK) between 2000-2014 with a complete UCLP and receiving treatment from nine UK cleft centres: West Midlands, North Thames, Evelina, South West, Spires, North West, Scotland, Leeds and Newcastle. For inclusion, children needed dental models to have been taken in infancy prior to primary surgical intervention and at the age of five years. The exposure variable was UCLP sidedness, and this was determined by two authors (L.E., B.R.) from 3-D scans of dental models of infants prior to their first surgical procedure. Patient records were used to confirm the complete UCLP cleft subtype and to identify the co-variable of patient sex.

### Outcome measure

The outcome variable was maxillary growth, assessed via occlusal relationships between the maxillary and mandibular arches on dental models taken at 5 years of age and analysed by the 5-Year-Old’s occlusal index described by Attack et al.^15^ The 5-Year-Old’s index has been accepted as the UK national audit tool at five years of age for the assessment of occlusal relationships in the primary dentition, prior to any orthodontic intervention.^16^

The 5-Year-Old’s index assigns a score of 1 (good) to 5 (poor) on an ordinal scale, based on the relationship of upper and lower dental model casts. Each cleft centre sent their dental models to be assessed by two nationally calibrated orthodontists, who scored the models individually. Differences were resolved through discussion to reach a consensus. All orthodontic examiners underwent a calibration process via the UK Orthodontic Clinical Excellence Network, with inter- and intra-rater kappa scores above 0.8, in order to be accepted as a scorer in this process.

### Analysis

The relationship between UCLP sidedness and maxillary growth was analysed initially using descriptive statistics and cumulative percentages. These associations were quantified by Odds Ratio (OR, 95%CI) and p values.^17^ Due to current contention about the best way in which to analyse ordinal data^18^, we took two regression approaches: First we calculated the OR of having a better outcome comparing left and right UCLP by logistic regression for each of the 4 potential cut points in the 5 Year-Old’s ordinal data. Each cut point dichotomised the 5-Year-Old’s index scores into better and worse outcome groups. If the common odds assumption was demonstrated to be satisfied, we used a proportional odds logistic regression to estimate a common odds ratio on sidedness effect.^19 20^ Adjustment was made for sex in each regression model in recognition of this potential explanatory variable in orofacial cleft sidedness research.^1^ Statistical analysis was performed using the R Project for Statistical Computing, version 4.3.1 (https://www.r-project.org)

## Results

There were 378 children included born with a complete UCLP, with data derived from nine UK cleft units. 256 (68%) had a right sided UCLP and 122 (32%) had a left sided UCLP. 248 (66%) were male and 130 (34%) were female. Five Year Old’s Index scores ranged from 1-5 with a mean score of 2.9 and a median score of 3.

Descriptive analysis (see Table 1 and Figure 1) shows a trend for slightly better maxillary growth in left sided UCLP, with a higher proportion of good 5-Year-Old’s Index Scores (1 and 2) in left sided UCLP (43%) compared to right sided UCLP (37%). Effect estimates in all regression models show a consistent trend for better growth in left sided UCLP with odds of having better outcome in left UCLP constantly higher than in right UCLP group (all ORs>1), yet there is weak evidence for a difference (as supported by the confidence intervals and p values) (see Table 2). The ORs comparing better to worse outcomes in left and right groups at different dichotomised cut points within the 5-Year-Old’s Index showed that the common odds assumption was reasonable (OR range between 1.02 to 1.33). The summary effect estimate from the proportional odds model describes the odds of having better 5-Year-Old’s Index Scores in left UCLP is 1.24 times higher compared to right UCLP having better 5-year-old’s index, (OR 1.24, 95% CI 0.85 to 1.82; p=0.27).

**Table 1.** 5-Year-Old’s (5YO) Index scores for left and right unilateral cleft lip and palate (UCLP) and females and males. Frequencies and cumulative percentages displayed.

| 5YO Index Score | Total | Left UCLP | Cumulative % | Right UCLP | Cumulative % | Female | Cumulative % | Male | Cumulative % |
| --- | --- | --- | --- | --- | --- | --- | --- | --- | --- |
| 1 | 44 | 31 | 12.1% | 13 | 10.7% | 12 | 9.2% | 32 | 12.9% |
| 2 | 112 | 80 | 43.4% | 32 | 36.9% | 36 | 36.9% | 76 | 43.5% |
| 3 | 97 | 65 | 68.8% | 32 | 63.1% | 32 | 61.5% | 65 | 69.8% |
| 4 | 79 | 49 | 87.9% | 30 | 87.7% | 29 | 83.8% | 50 | 89.9% |
| 5 | 46 | 31 | 100.0% | 15 | 100.0% | 21 | 100.0% | 25 | 100.0% |
| Total | 378 | 256 |  | 122 |  | 130 |  | 248 |  |

**Table 2:** Odds ratios of having better 5-Year-Old’s (5YO) Index scores comparing left to right sided unilateral cleft lip and palate with and without adjustment for sex.

| 5YO Index Analysis Strategy | Unadjusted OR (95% Cis) | P value | Adjusted OR (95% Cis) | P value |
| --- | --- | --- | --- | --- |
| Logistic regression:<br>Dichotomised cut points |  |  |  |  |
| • 1\|2 | 1.16<br>(0.58 to 2.29) | 0.68 | 1.18<br>(0.59 to 2.34) | 0.64 |
| • 2\|3 | 1.31<br>(0.84 to 2.04) | 0.23 | 1.33<br>(0.85 to 2.07) | 0.21 |
| • 3\|4 | 1.29<br>(0.82 to 2.02) | 0.28 | 1.31<br>(0.83 to 2.07) | 0.24 |
| • 4\|5 | 1.02<br>(0.53 to 1.97) | 0.96 | 1.05<br>(0.54 to 2.03) | 0.89 |
| Proportional odds model | 1.24<br>(0.85 to 1.82) | 0.27 | 1.27<br>(0.87 to 1.87) | 0.22 |

**Figure 1:**
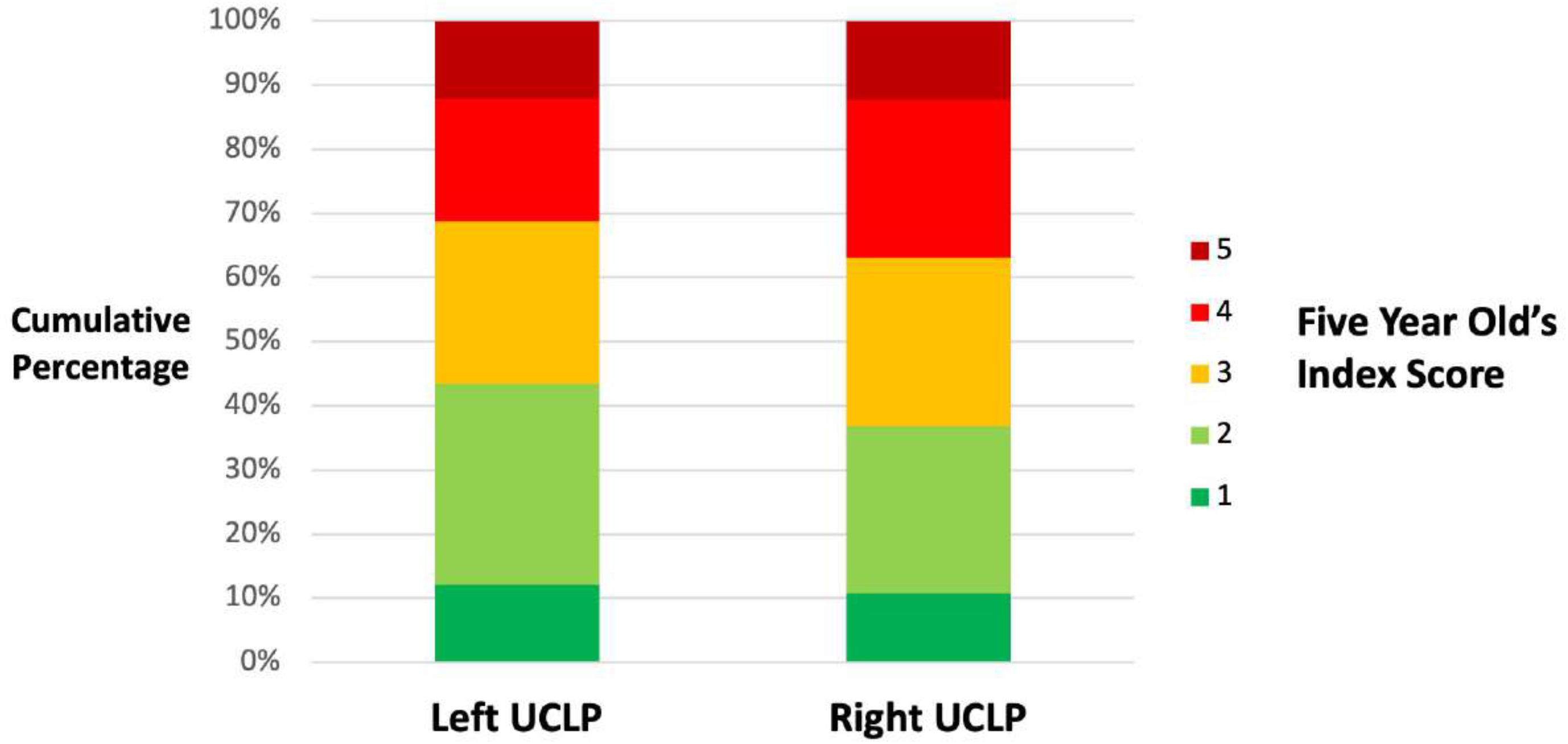
A cumulative bar chart to show the Five Year Old’s Index scores (Ordinal scale indicating maxillary growth from 1 (good) to 5 (poor)) for left and right sided unilateral cleft lip and palate (UCLP)

For the covariable of sex, there was a trend for better maxillary growth (5-Year-Old’s Index scores 1 and 2) in males (43.5%) compared to females (36.9%), although there was weak evidence in a univariate proportional odds regression analysis to suggest a difference (OR 1.42, 95% CI 0.97 to 2.08; p=0.07). Adjusting for sex in the sidedness regression models did not alter the trend of the unadjusted results.

## Discussion

### Summary of key findings

There was a trend for better maxillary growth in left compared to right sided UCLP at 5 years of age (OR 1.24, 95% CI 0.85 to 1.82; p=0.27), yet the uncertainty around the effect estimates in the regression analysis of ordinal 5-Year-Old’s Index scores implies the evidence for a difference in growth between left and right sided UCLP is weak.

### Strength and weaknesses

This study is strengthened by the sample of patients, which was heterogeneous in terms of the complete UCLP phenotype, and the consistent measurement of exposure and outcome variables. The inclusion of multiple UK cleft centres increases the applicability of the findings, yet introduces variability in terms of surgical techniques, timings and sequences, which are known to have an impact on maxillary growth.^21^ We adjusted for the co-variable of sex as this was readily available in our dataset but we were not able to add additional potentially important variables to our model, such as ethnicity.

The 5-Year-Old’s Index is an important outcome as it has been accepted as the recognised national audit measure in the UK and has been recommended for use in primary dentition.^16^ The long-term predictive ability of the 5-Year-Old’s Index has been questioned, particularly of poor growth (scores 4 and 5 on the 5-Year-Old’d Index), which has shown poor correlation with growth outcomes at skeletal maturity.^22^ The analysis of ordinal data in healthcare research is notoriously challenging and there is a paucity of a gold standard in terms of analytic strategies.^18^ Dichotomising the ordinal scale is appealing for simplicity yet the resulting effect estimate is only applicable for the arbitrarily chosen cut point and does not account for the variation of the data within each ordinal level.^19^ The use of the proportional odds logistic regression analysis maintains the integrity of the 5-Year-Old’s Index ordinal data and has been encouraged in healthcare outcome studies to retain clinically relevant information.^23^ The proportional odds model does rely on the assumption that the sidedness effect is constant across all cut points.^24^ This assumption was supported by our data with unadjusted odds ratios (Table 2) falling within a similar magnitude.

### Comparison to other studies

Whilst we are not aware of previous studies analyzing the relationship between UCLP sidedness and maxillary growth using the 5-Year-Old’s Index, other studies have investigated dental arch relationships using different outcome measures. Haque et al. ^25^ in a case series from Bangladesh reported no difference in dental arch relationships between left and right UCLP at a mean age of eight years (before orthodontic or orthognathic intervention) using the GOSLON yardstick, dichotomized into a binary outcome of favorable versus unfavorable growth (OR 1.12, 95%CI 0.409 to 3.11; p=0.83). Staudt et al.,^26^ in a case series from Switzerland reported better post-treatment occlusion at a mean age of 19 years for left sided UCLP using Modified Huddart Bodenham scores, although the prevalence of orthognathic surgery was higher for left UCLP (65% vs 20%). An Iranian study investigating cephalometric measurement of facial growth at an unspecified age, reported no difference in most of the 28 measurements reported between right and left UCLP (including vertical and sagittal skeletal, dental and soft tissue measures), but within the four measurements noted to be different there was greater mandibular length in left sided compared to right sided UCLP.^27^

### Interpretation and further work

The trend for better growth in left sided UCLP, reported in this study and previous studies, may suggest a subtle intrinsic influence of sidedness on maxillary growth outcomes. The severity of UCLP in terms of size of the alveolar cleft and circumference of the dental arch has been associated with maxillary projection.^11^ There is increasing evidence that sidedness may also be related to severity in the UCLP phenotype; right UCLP has been associated with a higher prevalence of additional congenital malformations^28,29^ and anatomical hypoplasia and asymmetries in the face.^2–4,30^ If patients born with right sided UCLP truly are at higher risk of maxillary growth disturbance, this has implications on patient counselling, and treatment options that might attempt to mitigate the increased risk.

The uncertainty around the effect estimate in this study may have a number of reasons. First, it is possible that UCLP sidedness does not impact maxillary growth. Second, the sample size may not have been sufficiently large enough to differentiate subtle differences in maxillary growth by sidedness. Third, the outcome measure at five years of age may not be the most appropriate to determine definitive growth outcome. Fourth, additional variables not considered within our model, both extrinsic (such as surgical technique) and intrinsic (such as ethnicity) may have an important influence. Further work is required to analyse maxillary growth in a large heterogenous sample of patients born with complete UCLP, ideally at skeletal maturity.

## Conclusion

We report a trend for better growth in left sided UCLP compared to right sided UCLP with a caveat that the strength of evidence for a difference is weak. Further work is warranted to investigate the impact of UCLP sidedness on outcomes including maxillary growth.

## Data Availability

All data produced in the present study are available upon reasonable request to the authors

## Notes

### Competing Interest Statement

The authors have declared no competing interest.

### Funding Statement

This study did not receive any funding

### Author Declarations

Ethical application assessed by the North West - Greater Manchester Central Research Ethics Committee (REC Ref: 18/NW/0536, IRAS: 225282, Research Registry UIN: 4068). Ethics approval was given.

